# An examination of the effect of dual task on gait variability in Parkinson’s disease and REM Sleep Behavior disorder

**DOI:** 10.64898/2026.05.22.26353152

**Authors:** Catherine L. Gallagher, Maureen B. Haebig, Ananya Heroor, Rachna Tiwari, David T. Plante, Ozioma Okonkwo, Jessica Baker, Laura Buyan-Dent, Teresa Mangin, Kathleen Shannon, Kristen A. Pickett

## Abstract

**Background:** Gait variability is a hallmark of Parkinson’s disease (PD) and has been linked to cognitive deficits and fall risk. Rapid eye movement sleep behavior disorder (RBD) is a strong predictor of synucleinopathies, yet evidence for gait changes in RBD is inconsistent. Performing a dual task increases gait variability, an effect that can be quantified using a cost function.

**Objective:** Determine the degree to which dual task cost differs between control, RBD, and PD participants at baseline, and between RBD converters versus non-converters at follow-up.

**Methods:** 46 RBD, 23 control, and 14 PD participants completed standardized gait analysis at baseline. Parameters chosen for analysis included enhanced gait variability index (eGVI), functional ambulation performance (FAP), velocity, step length, cadence, base of support, and double support time. Medical records were surveilled for 3 years following participant enrollment, determining that 6 RBD participants converted to PD or dementia. Baseline gait indices and dual task costs were compared between control, RBD, and PD groups at enrollment, and between RBD stable and RBD converters at follow-up.

**Results:** The PD group had greater eGVI, as well as greater dual task cost for FAP, cadence, width, and double support time. No differences in gait variability were identified between RBD and control groups at baseline. Compared to the stable group, RBD converters had greater dual task cost for FAP, velocity, cadence, and double support time.

**Conclusions:** Increased gait variability during dual task may identify RBD patients at imminent risk of phenoconversion.

## Introduction

Increased variability in walking is a symptom of many neurological disorders, including dementia and Parkinson’s disease (PD). Inability to maintain a rhythmical diagonal stepping pattern during distracting tasks has been described as a predictor of dementia and falls in community dwelling elderly populations (Ayers et al., 2014; Darweesh et al., 2019; Ma et al., 2022). From the standpoint of kinematic gait analysis, walking while performing an executive cognitive task produces an interference condition that tends to increase gait variability, especially in PD groups (Rochester et al., 2014; Uribe et al., 2022). Cognitive deficits in executive function and processing speed are typical of PD and impair gait automaticity (Snijders et al., 2016). Studies of the effect of dual task on kinematic features of gait in PD have shown an increase in gait variability during dual task performance (Raffegeau et al., 2019; Rochester et al., 2014).

REM sleep behavior disorder (RBD), a condition during which people act out dream behaviors during rapid eye movement (REM) sleep, is a strong risk factor for the future development of a synucleinopathy, especially Parkinson’s disease, Lewy body dementia (LBD), or multiple systems atrophy (MSA) (Postuma et al., 2019). Subtle increases in the variability of several kinematic gait parameters have been described in RBD (Ehgoetz Martens et al., 2019; McDade et al., 2013). The purpose of the present study was to examine the degree to which gait variability and changes in gait variability under dual task conditions distinguished RBD and PD from healthy controls and predicted clinical progression to Parkinsonism or dementia within the RBD cohort.

## Methods

### Participants

This study prospectively recruited persons with polysomnographically-proven REM sleep behavior disorder and demographically matched controls who had completed a sleep study that did not report RBD/REM sleep without atonia (RWA). Participants were 45 or more years of age, cognitively intact, and free of other major psychiatric, neurological, or poorly controlled medical illnesses, including moderate to severe sleep apnea. Case participants, the majority of whom also described dream enactment behavior, had REM sleep without atonia reported and in-house polysomnogram (PSG) by a sleep medicine physician. Apnea-hypopnea index needed to be lower than 15/hour or controlled as such on CPAP with persistent dream enactment behavior once obstructive sleep apnea was effectively treated. The control group consisted of a demographically matched sample that reported no history of dream enactment and had completed a polysomnogram reported as free of RWA/RBD. Data from a group of Parkinson’s PD participants enrolled in a pilot study, who had completed the same gait quantification protocol with the same equipment as the RBD and control groups, was included in the analysis. In addition to quantitative gait analysis, the research participants completed a structured interview, a neurocognitive evaluation, and brain MRI. The study was approved by the University of Wisconsin IRB (IRB # 2012-0037 and 2019-0987) and endorsed by the local VA R&D Committee.

### Procedures

All participants completed standardized cognitive screening and questionnaires that included the Geriatric Depression Scale (GDS) (Yesavage et al., 1982), Non-motor symptom questionnaire (NMSQ) (Chaudhuri et al., 2006), WRAT4 reading (Wilkinson and Robertson., 2006), Boston naming test (Kaplan et al., 1983), category and phonemic fluency (Gladsjo et al., 1999), Trail Making Tests A and B (Reitan, 1992), Stroop (Golden, 1975), Wisconsin Card Sort-64 (Paolo et al., 1996), Hooper Visual Organization Task (Merten, 2004), Judgement of Line Orientation (Calamia et al., 2011), and Rey Auditory Verbal Learning Task (RAVLT) (Rey, 1964). PD participants completed the Hopkins Verbal Learning Test (HVLT) (Benedict et al., 1998) instead of the RAVLT. A trained movement disorders neurologist completed the MDS-UPDRS III (Goetz et al., 2008) for control/RBD participants and the UPDRS III (Fahn, S. and Elton, R., 1987) for PD participants. Although the MDS-UPDRS is a more detailed scale, scores from the newer and older versions of the scale are generally correlated (Tosin et al., 2021). They also completed a non-motor symptom questionnaire (NMSQ) (Chaudhuri et al., 2006) and the University of Pennsylvania Smell Identification Test (UPSIT) (Doty et al., 1984).

Gait assessment was completed on a Zeno walkway (Protokinetics LLC, Havertown, PA). Each participant completed five self-paced passes over the walkway while performing each of five tasks, administered in random order: 1) forward walking at preferred speed, 2) forward walking at a fast speed, 3) forward walking while performing a dual task, 4) forward tandem walking, and 5) backward walking at preferred speed. A start/finish line was located one meter from each end of the mat to allow for capture of steady state walking without acceleration or deceleration. Participants were closely monitored during all trials to prevent falls and injury. If the individual left the walkway prior to completion of the task, stumbled, or needed assistance from the spotter, the trial was discarded and immediately repeated. Dual task walking consisted of preferred-speed forward walking while verbally listing as many items as possible in a given category (animals, colors, games with a ball, things in the sky, foods), with number of correct, unique, responses recorded. Gait data were collected in two different laboratories due to the first laboratory being closed for construction. The second location differed from the first in having sufficient but less extensive space surrounding the mat, and having 12×12 floor tiles of muted but alternating colors. The same gait mat was used in both spaces and locations were selected to minimize superfluous noise and distractions. Primary gait outcomes were selected a priori to represent pace, rhythm, spatial control, stability, variability, and global gait performance.

Medical records were surveilled for 3 years following participant enrollment and baseline testing. Those persons who converted to Parkinsonism or Lewy body dementia were termed “converted” and their baseline data compared with non-converters and controls. Data from those participants who died or were lost to follow-up was designated as “missing” for the purposes of conversion to Parkinsonism; i.e. baseline data from those RBD participants who were known to be healthy at follow-up were compared with those who converted.

### Data analysis

Demographic data and normalized neuropsychometric test scores were compared across groups (Control, RBD, and PD) via ANOVA. Standard gait outputs from the walking trials, generated from macros by the PKMAS software version 509c3, were analyzed in SPSS (IBM, Version 31.0, Chicago). For each gait condition, means were calculated across all valid steps for step length (cm), velocity (sum of stride lengths/sum of stride times; cm/sec), cadence (steps/min), width/base of support (cm), and double support time (period within each gait cycle when both feet are in contact with the mat; msec). Summary measures of gait variability (enhanced gait variability index; eGVI) (Gouelle et al., 2018), and functional ambulation performance (FAP) (Gretz et al., 1998) were also evaluated. To provide a normalized measure of the effect of dual task on gait function, we then calculated dual task cost (normal walking – dual task walking)/normal walking *100. Each gait variable of interest was analyzed via ANCOVA, in which the independent variable was the gait parameter, fixed factor was the participant group (control, RBD, PD), and covariates included age and testing location. Group differences were described as significant based on a pre-determined threshold of *P* < 0.05.

For the longitudinal analysis, the Welch’s t-test (pre-selected 2-sided *P*<.05) was used to compare baseline gait parameters between RBD participants who converted to Parkinsonism or Lewy body dementia within the follow-up period, to those who were known not to convert. The Welch’s test (Welch, 1947) is an adaptation of the Student’s t-test that is more reliable in the case of unequal variances or sample sizes. Then, because the groups of converters versus nonconverters differed by age, the unpaired test was repeated after all persons younger than age 60 at baseline visit had been removed from the non-converter group.

## Results

Mean values for demographics, questionnaires, and normalized psychometric assessments are shown in Table 1. There were no significant differences in age, years of formal education, or neuropsychological performance scores between control, RBD, and PD participant groups at baseline. The RBD and control groups had a higher proportion of males than the PD group.

**Table 1:**
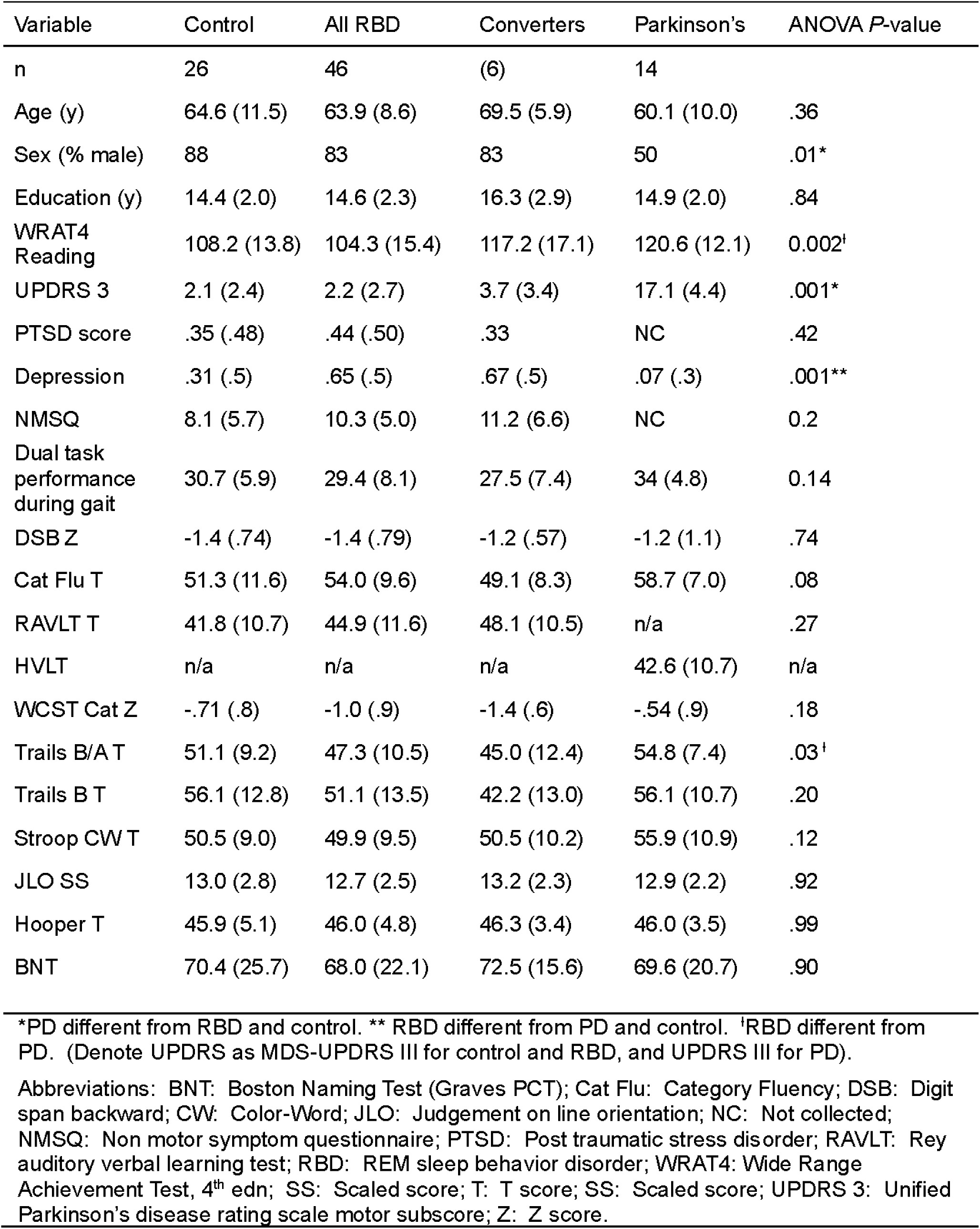
Baseline demographic and psychometric data (mean/SD)

UPDRS motor sub scores were higher, as expected, for the PD group, than MDS-UPDRS motor sub scores for the control and RBD groups. The RBD group endorsed a greater burden of depressive symptoms in comparison to the PD and control groups. A higher incidence of depression and anti-depressant therapy has been noted previously in RBD cohorts (McDade et al., 2013).

Of the sample, 12 participants (4 control, 8 RBD) completed gait testing in both laboratory locations. There was an increase in eGVI, as well as a reduction in velocity, step length, and cadence in the second location. Therefore, gait location was entered as a covariate into the ANCOVA analyses. Of note, all of the Parkinson’s participants completed testing in the second lab location.

Within group analysis showed a main effect of gait task between normal and dual task gait for all three participant groups, such that participants walked slower, spent more time in double support, demonstrated greater eGVI and decreased FAP (p<0.01 for all) during dual task.

Individuals with PD did not significantly shorten step length from normal to dual task (p = 0.056), whereas control (p = 0.014) and RBD (p = 0.023) groups took shorter steps. However, the PD group did walk with a wider base of support during dual task gait (p = 0.01), while the control (p = 0.884) and RBD (p = 0.621) groups did not significantly alter their step width. Mean scores for each group and condition are shown in Table 2.

**Table 2:** Mean of gait parameters and dual task costs for participant groups.

| Variable | Trial | Control | RBD | Converters | Parkinson's | ANCOVA |
| --- | --- | --- | --- | --- | --- | --- |
| n |  | 26 | 46 | 6 | 14 |  |
|  |  | Mean (SD) | Mean (SD) | Mean (SD) | Mean (SD) | P value |
| eGVI | Normal | 119.7 (13.9) | 115.8 (11.5) | 114.3 (9.5) | 124.3 (8.9) | .016* |
|  | Dual | 130.6 (12.2) | 130.9 (15.3) | 134.0 (16.1) | 143.0 (13.8) | .003*/.007** |
|  | Cost (%) | 10.1 (11.8) | 13.3 (12.9) | 17.7 (15.4) | 15.4 (11.7) |  |
| FAP | Normal | 95.9 (3.5) | 92.8 (9.6) | 98.6 (2.1) | 93.1 (6.4) |  |
|  | Dual | 85.9 (8.4) | 85.4 (13.1) | 83.6 (7.5) | 75.6 (17.7) |  |
|  | Cost (%) | 10.4 (8.9) | 7.6 (8.9) | 15.2 (7.5) | 19.2 (16.9) | .031* |
| Velocity (cm/sec) | Normal | 101.9 (15.0) | 101.1 (22.4) | 102.5 (8.2) | 103.4 (23.7) |  |
|  | Dual | 82.9 (15.9) | 85.8 (21.6) | 74.5 (11.4) | 77.0 (38.1) |  |
|  | Cost (%) | 17.3 (3.4) | 17.9 (2.6) | 27.2 (10.0) | 26.9 (7.0) |  |
| Step length (cm) | Normal | 61.5 (7.1) | 61.9 (8.5) | 63.3 (2.9) | 62.8 (12.4) |  |
|  | Dual | 58.4 (7.1) | 59.8 (8.9) | 57.4 (4.9) | 57.5 (16.1) |  |
|  | Cost (%) | 4.58 (9.54) | 2.94 (9.45) | 9.26 (7.23) | 9.01 (15) |  |
| Cadence (steps/min) | Normal | 98.8 (7.8) | 96.9 (13.4) | 96.8 (5.4) | 99.1 (13.4) |  |
|  | Dual | 84.7 (11.2) | 85.3 (11.9) | 77.5 (8.2) | 77.1 (19.7) |  |
|  | Cost (%) | 14.0 (11.4) | 11.7 (10.9) | 19.9 (7.3) | 22.3 (16.3) | .047* |
| Width (cm) | Normal | 10.2 (3.4) | 9.5 (3.9) | 10.3 (2.8) | 9.7 (4.4) |  |
|  | Dual | 10.1 (4.8) | 9.8 (4.8) | 9.4 (3.6) | 11.9 (5.1) |  |
|  | Cost (%) | -2.3 (26.7) | 2.7 (26.7) | -8.0 (23.9) | 26.5 (36.6) | .045* |
| Double Support Time (msec) | Normal | 394.4 (63.9) | 399.8 (110.4) | 379.5 ((55.3) | 374.3 (92.3) |  |
|  | Dual | 492.5 (100.8) | 473.7 (139.2) | 530.8 (86.0) | 635.6 (352.4) |  |
|  | Cost (%) | 25.7 (22.6) | 20.5 (25.8) | 40.8 (19.8) | 70.1 (88.0) | .056**/.017* |

Abbreviations: eGVI, enhanced gait variability index; FAP, functional ambulation performance index.
ANCOVA: Comparing Control, RBD, and PD at baseline; converters are a subgroup of RBD.
\*Post hoc P value for PD versus RBD; \*\*PD versus Control

Comparison of gait parameters between groups at baseline (Table 2) demonstrated that the PD group had greater gait variability (eGVI) under both normal and dual task walking conditions than RBD and control participants. Parkinson’s participants also showed greater dual task cost for functional ambulation performance (FAP), cadence, width, and double support time. For several measures (FAP, step length, cadence, double support time), the lowest dual task cost was seen for the RBD group rather than the control group. With regard to width/base of support, the control and RBD groups made little adjustment of stride width during dual task, while PD participants widened their base of support by 26%. RBD converters paradoxically adopted a narrower mean base of support during dual task. There were no significant differences between the RBD group and the control group at baseline. However, we did note that for the RBD converter group, dual task cost for most parameters (eGVI, FAP, velocity, step length, cadence, double support time) was higher and more similar to the PD group than to the control group. To illustrate this point, data from the converter group (a subgroup of the RBD group) is included in Table 2.

A comparison of baseline demographic and psychometric data from RBD participants for whom follow-up data was available (36 stable RBD, 6 converters; Table 3) showed that converters were older (mean age 69.50+/− 6.0 years) than non-converters (62.31+/−8.4 years). Years of formal education and neuropsychometric assessment scores did not differ significantly between converters and stable RBD participants, although there was a trend towards lower Trails B T scores and higher Boston Naming test normalized scores in converters (p=.07). In comparison to non-converters, converters had no significant difference in MDS-UPDRS motor subscores, NMSQ scores, nor exposures to smoking, caffeine, and Agent Orange. They did have a higher incidence of reported urinary dysfunction, erectile dysfunction, diabetes, and olfactory loss based on the NMSQ (Chaudhuri et al., 2006). All 6 who converted to neurodegenerative disease had olfactory loss.

**Table 3:** Comparison of baseline data for RBD groups.

| Variable | RBD stable | RBD stable (age-matched) | RBD converters | 2 Tailed P-value (36/23 vs 6) |
| --- | --- | --- | --- | --- |
| n | 36 | 23 | 6 |  |
| Age (y) | 62.3(8.4) | 67.7 (4.7) | 69.5 (5.9) | .03*/.5 |
| Sex (% male) | 80 | 87 | 83 | .9/.8 |
| Education (y) | 14.4 (2.9) | 14.4 (2.1) | 16.3 (2.9) | .2/.2 |
| UPDRS3 | 1.8 (2.6) | 1.8 (1.3) | 3.7 (3.4) | .3/.2 |
| PTSD score | .46 (.5) | .43 (.5) | 0.33 | .6/.7 |
| Depression | .65 (.5) | .6 (.5) | .67 (.5) | .8/.7 |
| NMSQ | 9.9 (4.6) | 10.4 (4.3) | 11.2 (6.6) | .7/.8 |
| DSB Z | -1.5 (.8) | -1.7 (.7) | -1.2 (.57) | .3/.13 |
| Cat Flu T | 54.6 (10.2) | 55.5 (11) | 49.1 (8.3) | .5/.2 |
| RAVLT T | 44.5 (12.1) | 45.7 (13.3) | 48.1 (10.5) | .3/.6 |
| WCST Cat Z | -.9 (1.0) | -1.2 (.9) | -1.4 (.6) | .13/.7 |
| Trails B/A T | 48.7 (10.0) | 48.1 (10.7) | 45.0 (12.4) | .5/.6 |
| Trails B T score | 54.4(12.3) | 54.2 (12.8) | 42.2 (13.0) | .07/.8 |
| Stroop CW T | 50.1 (9.8) | 49.3 (10.5) | 50.5 (10.2) | .9/.8 |
| JLO SS | 12.9 (2.4) | 12.8 (2.6) | 13.2 (2.3) | .8/.7 |
| Hooper T | 45.7 (5.0) | 45.0 (4.4) | 46.3 (3.4) | .7/.4 |
| BNT (Graves) | 66.2 (22.8) | 63.8 (22.2) | 72.5 (15.6) | .08/.06 |

\*RBD stable (36) different from converters by 2-tailed T test
Abbreviations: BNT: Boston Naming Test (Graves PCT); Cat Flu: Category Fluency; DSB: Digit span backward; CW: Color-Word; JLO: Judgement on line orientation; NMSQ: Non motor symptom questionnaire; PTSD: Post traumatic stress disorder; RAVLT: Rey auditory verbal learning test; SS: Scaled score; T: T score; SS: Scaled score; Z: Z score.

Bar graphs showing mean dual task cost for velocity, step length, cadence, and total double support, collected prior to disease conversion, are shown in Figure 1. This figure is included to demonstrate the similarity of dual task impact between PD and RBD converter groups in comparison to RBD stable and control groups. Baseline gait data from stable RBD participants versus RBD converters is also shown in Table 4. Compared to the stable group, RBD converters had greater dual task cost for FAP, velocity, cadence, and double support time.

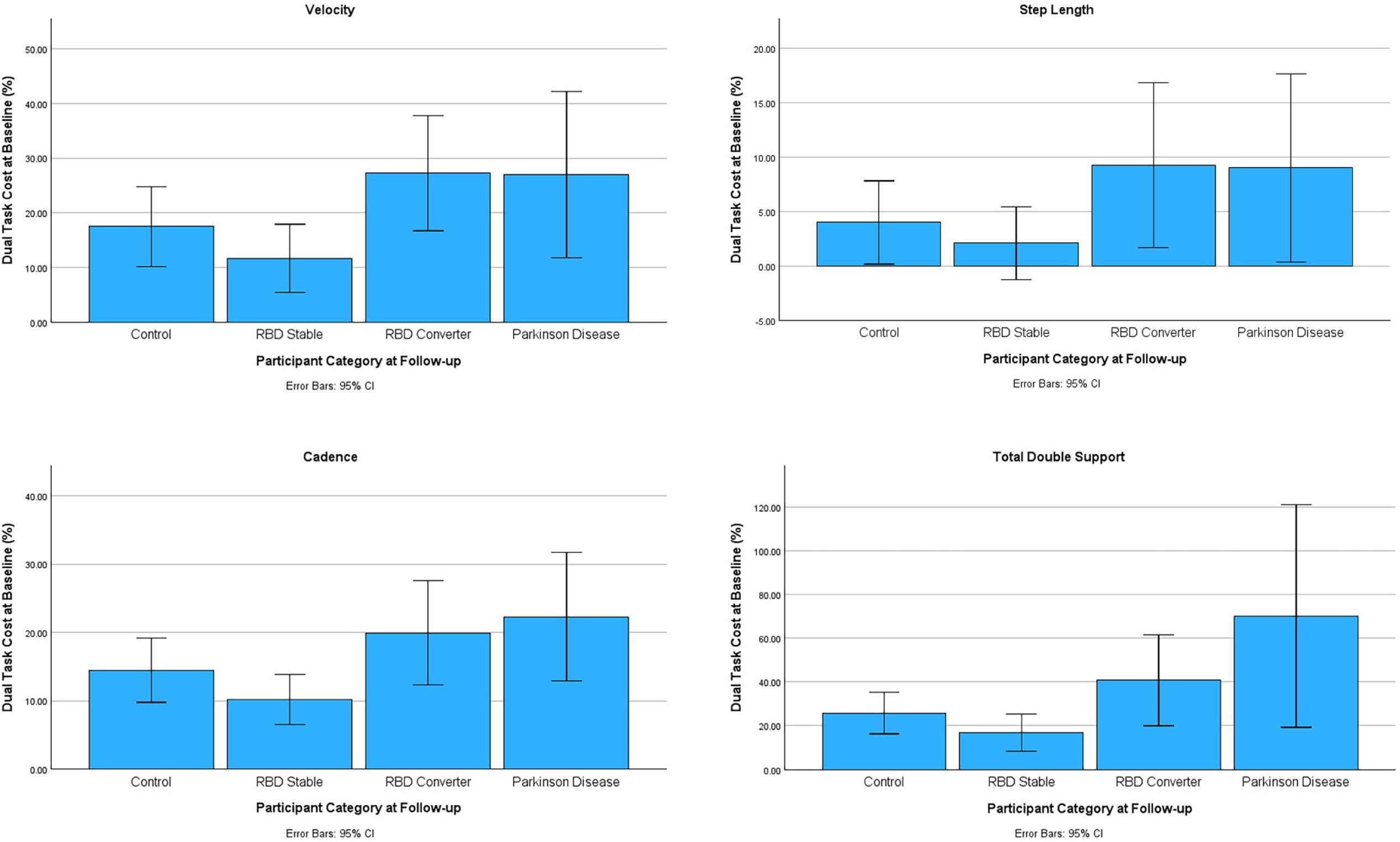

**Table 4:** Mean gait parameters and costs for RBD sub groups.

| Variable | Trial | RBD stable | RBD stable (age-matched) | RBD converters |  |
| --- | --- | --- | --- | --- | --- |
| n |  | 36 | 23 | 6 |  |
|  |  | Mean (SD) | Mean (SD) | Mean (SD) | *P* value |
| eGVI | Normal | 115.4 (12.0) | 114.9 (12.8) | 114.3 (9.5) |  |
|  | Dual | 130.2 (15.0) | 132.3 (12.7) | 134.0 (16.1) |  |
|  | Cost (%) | 13.3 (12.8) | 15.9 (13.0) | 17.7 (15.4) |  |
| FAP | Normal | 93.1 (9.7) | 93.3 (10.2) | 98.6 (2.1) | .006*/.03** |
|  | Dual | 87.1 (12.9) | 84.4 (13.2) | 83.6 (7.5) |  |
|  | Cost (%) | 5.9 (14.0) | 9.1 (14.1) | 15.2 (7.5) | .05* |
| Velocity (cm/sec) | Normal | 103.6 (23.1) | 101.2 (22.6) | 102.5 (8.2) |  |
|  | Dual | 90.0 (21.7) | 87.5 (23.7) | 74.5 (11.4) | .022*/.07** |
|  | Cost (%) | 11.6 (18.5) | 12.4 (18.4) | 27.2 (10.0) | .01*/.02** |
| Step length (cm) | Normal | 62.8 (8.5) | 62.4 (9.2) | 63.3 (2.9) |  |
|  | Dual | 61.3 (9.1) | 60.7 (10.0) | 57.4 (4.9) |  |
|  | Cost (%) | 2.1 (9.9) | 2.4 (9.6) | 9.3 (7.2) | .06*/.08** |
| Cadence (steps/min) | Normal | 97.7 (13.8) | 96.3 (13.3) | 96.8 (5.4) |  |
|  | Dual | 87.4 (13.7) | 85.8 (14.9) | 77.5 (8.2) | .03*/.09** |
|  | Cost (%) | 10.2 (10.9) | 10.6 (11.5) | 19.9 (7.3) | .02*/.03** |
| Width (cm) | Normal | 9.4 (4.0) | 9.1 (4.2) | 10.3 (2.8) |  |
|  | Dual | 9.5 (4.8) | 9.4 (5.2) | 9.4 (3.6) |  |
|  | Cost (%) | 2.9 (31.2) | 3.9 (36.7) | -8.0 (23.9) |  |
| Double Support Time (msec) | Normal | 398.22(117.0) | 417.1 (120.4) | 379.5 (55.4) |  |
|  | Dual | 456.6 (142.6) | 485.4 (146.5) | 530.83 (86.0) |  |
|  | Cost (%) | 16.8 (25.3) | 18.73 (27.1) | 40.8 (19.8) | .03*/.048** |

Abbreviations: eGVI, enhanced gait variability index; FAP, functional ambulation performance index.
\*RBD versus converters; \*\*Age-matched RBD versus converters.

However, the converter group had an unusually high FAP at baseline, which likely influenced FAP cost. Repeating the comparison using data from stable RBD participants of age 60 years or more (23, mean age 67.7) confirmed higher dual task cost for velocity, cadence, and double support time for converters compared with this age-matched group.

## Discussion

Gait variability is a hallmark of Parkinson’s disease and has been linked to cognitive deficits and fall risk. Rapid eye movement (REM) sleep behavior disorder (RBD) is a strong predictor of synucleinopathies, yet evidence for gait changes in RBD is inconsistent. This study examined the degree to which gait variability and changes in gait variability under dual task conditions distinguished RBD and PD from healthy controls and predicted clinical progression to Parkinsonism or dementia within the RBD cohort. Dual-task gait variability, particularly reductions in cadence, velocity, and increases in double support, may serve as a biomarker for imminent conversion in RBD. The findings also support eGVI as a valid measure of gait variability in PD.

Whereas two previous studies (Ehgoetz Martens et al., 2019; McDade et al., 2013) have shown RBD patients to have subtly increased variability in kinematic features of gait, the present study did not replicate these findings, even under dual task conditions. However, the results of the two prior studies were not necessarily consistent with one another. The first (McDade et al.) showed probable RBD participants to have decreased velocity and cadence, as well as increased variability in double limb support, stride time, and swing time, compared with controls, during a single self-paced 10-meter walking trial of two passes over the mat (McDade et al., 2013). This study enrolled a similar-sized RBD group (42) to the present study, but a much larger control group (492 individuals), thus increasing statistical power to detect subtle differences. While the present study used polysomnographic evidence of RWA to confirm RBD, the former (McDade et al) study used a single-question screen that has been validated against polysomnogram as sensitive for RBD/RWA (Rochester et al., 2014). The second study (Ehgoetz et al.), of 24 RBD participants and 12 controls, showed no group differences during normal walking, but that the RBD failed to widen their base of support, and showed increased step width variability during dual task (Ehgoetz Martens et al., 2019). In the present study, RBD non-converters made a normal width adjustment during dual task, whereas the converter group narrowed their base, which replicates findings from the previous study, but only amongst imminent converters.

In the present study, the PSG-proven RBD participants showed modestly lower gait variability (eGVI) scores during normal walking, as well as lower dual task cost for FAP, step length, cadence, and double support, in comparison to the control group. It is possible that the RBD group, although matching the control group for basic demographic factors and recruitment source, was physiologically healthier. This conclusion is not supported by the cognitive data (Table 1), which shows fairly matched scores between controls and RBD, but could explain why we failed to detect an above normal gait variability in the full RBD group (46 participants) in comparison to controls at baseline. Thus, differences in study populations, gait analysis methodology, and statistical power, likely explain our inability to replicate findings from these previous studies.

During this study, the incidence of conversion to a neurodegenerative disease within the RBD cohort was approximately as expected: 6/36, or 17%, over 3 years. The average conversion rate in RBD has been estimated at 6.3% per year (McDade et al., 2013). When we compared baseline gait data from the six RBD participants who developed a neurodegenerative disorder during the follow-up interval, to those who remained stable, we found the gait profile of converters to be similar to Parkinson’s participants; in spite of low statistical power for the small converter group, this difference included a significant reduction in velocity and cadence, as well as an increase in double support, during dual task. These results suggest that changes in the kinematic features of gait may be useful as biomarkers to predict conversion to Parkinsonism or dementia within RBD cohorts. Additional potential biomarkers in the converter group included a higher incidence of diabetes, genitourinary dysautonomic symptoms, and olfactory loss, as well as a trend towards lower T scores on Trails B.

In the present study, we did observe that data from a relatively small group of Parkinson’s participants showed greater eGVI than control and RBD groups during both normal and dual task walking. Since a large number of variables are produced during standardized gait analysis, the Gait Variability Index (GVI), which incorporates step to step differences in eight parameters, was developed as a summary measure for variability. However, GVI scores for PD individuals were found to fall close to the normal range, and to discriminate poorly between PD Hoehn and Yahr (HY) stages 2 and 3 (Snjders et al., 2016). Limitations in the GVI were subsequently addressed through development of the enhanced gait variability index (eGVI), which reduced the redundancy of variables (using 5 variables of step length, step time, stance time, single support time, and stride velocity), and addressed magnitude and directionality problems (Zhou et al., 2025). The eGVI uses 1 + the natural log of net difference between successive spatiotemporal values to generate an index for which 100 signifies a healthy population mean, and a 10-point difference corresponds to 1 SD. An eGVI above 100 indicates gait variability above the mean and less than 100, below the mean (Zhou et al., 2025). A study of eGVI in 273 patients with Parkinson’s disease showed that patients with monthly to daily falls have a mean eGIV of ∼120 or higher, in comparison to rare fallers, and that eGVI explains approximately 15% of the variance in fall frequency; however, this study did not include a healthy control group (Raffegeau et al., 2019). In healthy community dwelling individuals, eGVI has been shown to correlate with visuospatial and delayed memory function (Postuma et al., 2019). Thus, our finding that eGVI was significantly different in a relatively small group of Parkinson’s patients is novel and suggests that it is a valid measure of gait variability in PD. Since all of the Parkinson’s patients were studied in a location with variable floor tiles outside the mat, it is possible that two stressors (both visual stimuli and dual task) enhanced the detection of gait variability in PD.

## Conclusions

This study had several limitations: A small control group compared with some prior studies; higher gait variability amongst the control group in comparison to the RBD group during normal walking that may have limited our ability to detect increased gait variability during dual task; and change in lab location during the study with Parkinson’s participants only completing the gait analysis in the second location. Nonetheless, the results of this study suggest that gait variability increases immediately prior to the development of Parkinsonism or dementia in RBD and as such may be a useful biomarker for imminent phenoconversion.

## Data Availability

All data were collected at the Department of Veterans Affairs and the signed subject consent forms and HIPAA authorization did not include provisions for making the individual data publicly available. However, the authors may be able to provide metadata - i.e. the numerical, deidentified, data, subject to approval from VA research oversight committees and privacy officer. Requests for access can be sent to: Lynn Tarpey, Privacy Officer, William S. Middleton Memorial Veterans Hospital, 2500 Overlook Terrace Dr., Madison, WI, 53705, PH: 608-256-1901.

## Notes

### Competing Interest Statement

The authors have declared no competing interest.

### Funding Statement

This work was supported by VA Merit Review award number I01CX000555-6-9 from the United States (U.S.) Department of Veterans Affairs Clinical Sciences Research and Development Service. The contents do not represent the views of the U.S. Department of Veterans Affairs or the United States Government. Material support including data transfer and storage, as well as advice on analysis techniques was provided by the Wisconsin Alzheimer's Disease Research Center (NIH 2P30AG062715).

### Author Declarations

Ethical considerations and consent to participate: Data used in this report were acquired during two human subjects studies that were approved by the University of Wisconsin IRB (IRB # 2012-0037 and 2019-0987) and endorsed by the local VA Research and Development Committee. Informed written consent was provided by all study participants prior to study procedures.

